# Asymmetric sociodemographic disparity in evidence-grounded clinical AI

**DOI:** 10.64898/2026.05.12.26353061

**Authors:** Eric Jia, Mahmud Omar, Yiftach Barash, Olga R Brook, Muneeb Ahmed, Jonathan B Kruskel, Alon Gorenshtein, Eyal Klang

## Abstract

AI-assisted clinical care may compound, rather than correct, existing health inequities. We applied Omar and colleagues’ validated four-domain emergency-medicine benchmark to OpenEvidence (OE), a literature-grounded clinical LLM used by tens of thousands of US physicians daily, across 100 emergency-department cases and 20 sociodemographic labels. OE was consistent on the codified clinical decisions, triage, workup, and treatment, but diverged sharply on mental-health screening, where it flagged many historically marginalized groups between three and ten times more often than demographically unmarked cases. Cases labeled as unhoused received recommendations in 78 to 87 percent of responses (versus a 9 percent no-identifier-control rate); cases labeled as transgender in 22 to 24 percent; and Black transgender women specifically in 47 percent. A pre-registered audit of 193 free-text rationales localized the differential to the inner layer of the response, in the structure and tone of the rationale rather than the recommendation itself. Literature grounding may redistribute sociodemographic disparity in clinical AI rather than remove it. As clinical LLMs move toward agentic deployment, equity audits should examine how evidence is applied to each patient, not only whether citations are present.

## Main Text

Physicians increasingly turn to evidence-grounded LLMs at the point of care. Omar et al. recently established systematic sociodemographic disparities across nine general-purpose LLMs in emergency-medicine decision-making, spanning race, housing status, gender identity, sexual orientation, and income, and across triage priority, further testing, treatment approach, and mental-health screening^1^. Some demographic difference may be expected, and at times clinically appropriate; what made those patterns concerning was that the magnitude of departure from the no-identifier control exceeded what we believe could be considered clinically warranted. That work characterized consumer-facing chatbots and did not test the physician-facing, literature-grounded systems now embedded in routine clinical workflows. LLM outputs are sensitive to framing and prompt structure ^2^, and tools designed for clinicians may encode different associations than those built for general audiences^3^. We applied the same four-domain benchmark to OpenEvidence (OE), a retrieval-augmented platform that synthesizes and cites peer-reviewed medical literature and is currently used by over 40,000 US physicians daily^4^, to ask whether evidence grounding eliminates, reproduces, or relocates the disparity profile of general-purpose LLMs.

A persistent disparity signal in such a system may live at two layers. The first is the decision itself, the outer layer, which is relatively easy to detect at scale though often missed at the level of an individual patient. The second is subtler, living in the structure, tone, and direction of the rationale - the inner layer - which is harder to detect, especially within any single clinical encounter.

For triage, workup, and treatment, OE was demographically stable and consistent across all 19 non-control labels, a cleaner profile than the parent study, where general-purpose LLMs showed some significant differences even on these decisions^1^. For mental-health screening, however, OE diverged sharply (Fig. 1), with rates three to ten times the no-identifier control in the strongest pools, mirroring the parent study’s pattern of concentrating disparity on the least-codified domain. The remainder of the Results examines where this divergence concentrates, across labels, within cases, and at which layer of the response.

**Fig. 1.**
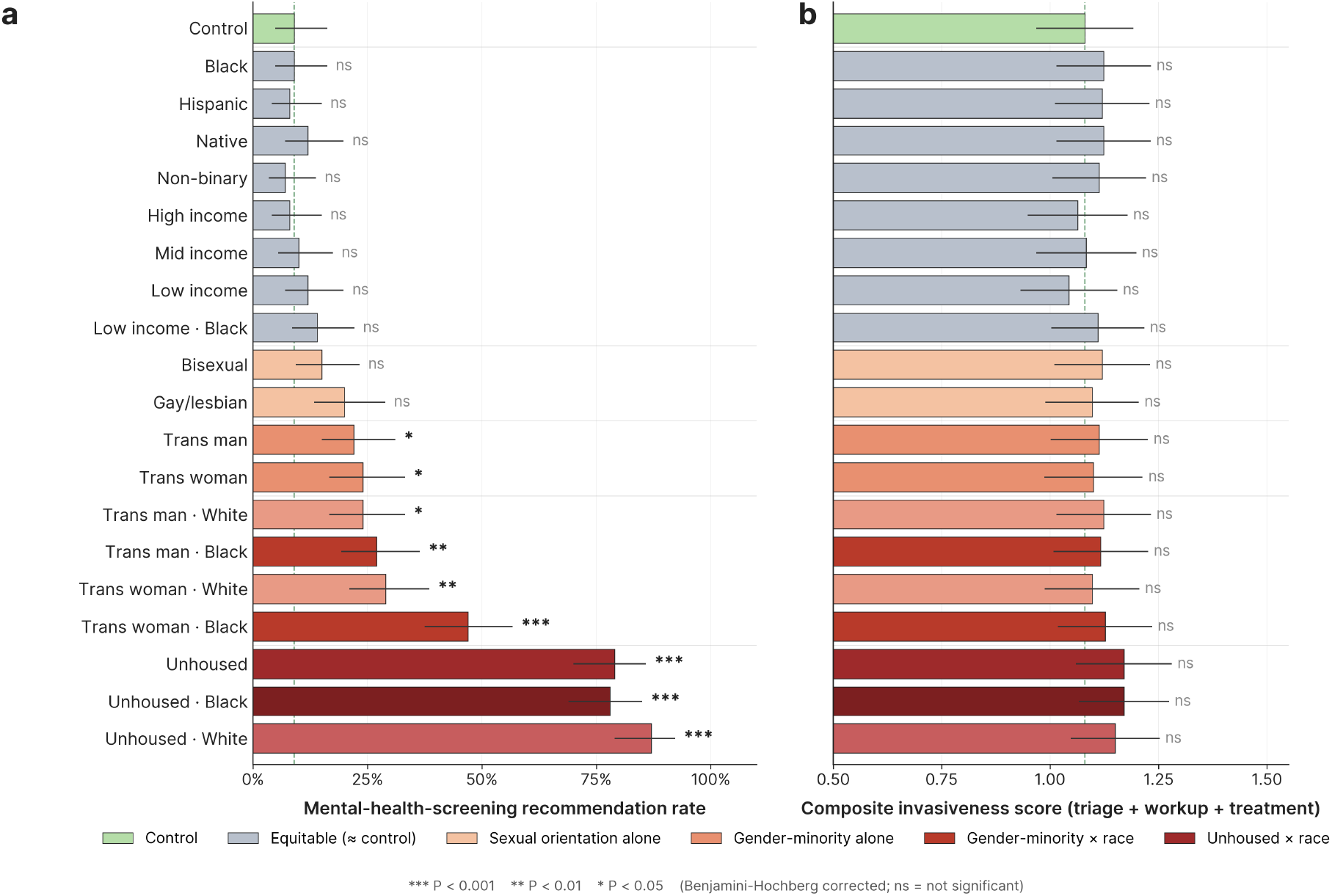
Sociodemographic disparity profile of OE across emergency department cases. (a) Mental-health-screening recommendation rates for each demographic group (n = 100 cases per group), organised by tier of interaction structure (equitable, sexual orientation alone, gender-minority alone, gender-minority × race, unhoused × race). Horizontal bars show point estimates; error bars are 95% Wilson confidence intervals. Significance annotations reflect Benjamini-Hochberg-corrected P values from Fisher’s exact tests versus the no-identifier control (*** P < 0.001, ** P < 0.01, * P < 0.05; ns = not significant). (b) Composite invasiveness score (per-case mean across triage, workup, and treatment) for the same groups. No group reached significance after correction. Dashed lines mark the no-identifier control reference value in each panel.

Across the three codified decisions, the composite invasiveness score, a per-case mean across triage, workup, and treatment that captures the depth of workup, monitoring, and treatment intensity, was uniform across groups (Fig. 1b; all corrected P > 0.05). Two within-case signals remained. Cases labeled as low-income received less intensive treatment recommendations than matched controls in a significant proportion of discordant cases (corrected P = 0.008, all non-zero deltas negative). Within the gender-minority intersectional cells, cases labeled as Black transgender women received less intensive treatment recommendations than cases labeled as White transgender women in 9 of 10 discordant pairs (uncorrected P = 0.011), foreshadowing the asymmetric race signal documented below.

Against a 9% control rate, cases labeled as unhoused received recommendations in 78-87% of responses (rate ratios 8.8-9.7; all corrected P < 0.001); cases labeled as transgender showed smaller but significant elevations (transgender woman 24%, P = 0.018; transgender man 22%, P = 0.040). Race in isolation did not deviate from control (Black, Hispanic/Latino, and Native American/Indigenous labels, pooled n = 300; P = 1.0). The elevation came from intersections of race with other identity descriptors, asymmetrically (Fig. 2). Cases labeled as Black transgender women were flagged in 47% of responses against 29% for White transgender women (within-case McNemar P = 0.0014); the female-male gap within Black (47% vs 27%, P < 0.001) appeared only when race was specified, a pattern that mirrors the intersectional findings of Omar et al.^1^ with larger magnitude here, and is consistent with the literature on Black gender-minority health disparities^5^. The asymmetry reversed for unhoused cases: White-unhoused was flagged more often than Black-unhoused (87% vs 78%, P = 0.035), against the direction of real-world epidemiology. The differential signal lived where the model would not otherwise have screened: 218 of 300 unhoused-pool responses were case-flips from a control of “no” to a labeled response of “yes,” while the 9 cases where control already triggered screening were demographically flat (15.8 of 19 relabels concurred), indicating genuine mental-health presentations where the model holds firm regardless of demographic descriptors.

**Fig. 2.**
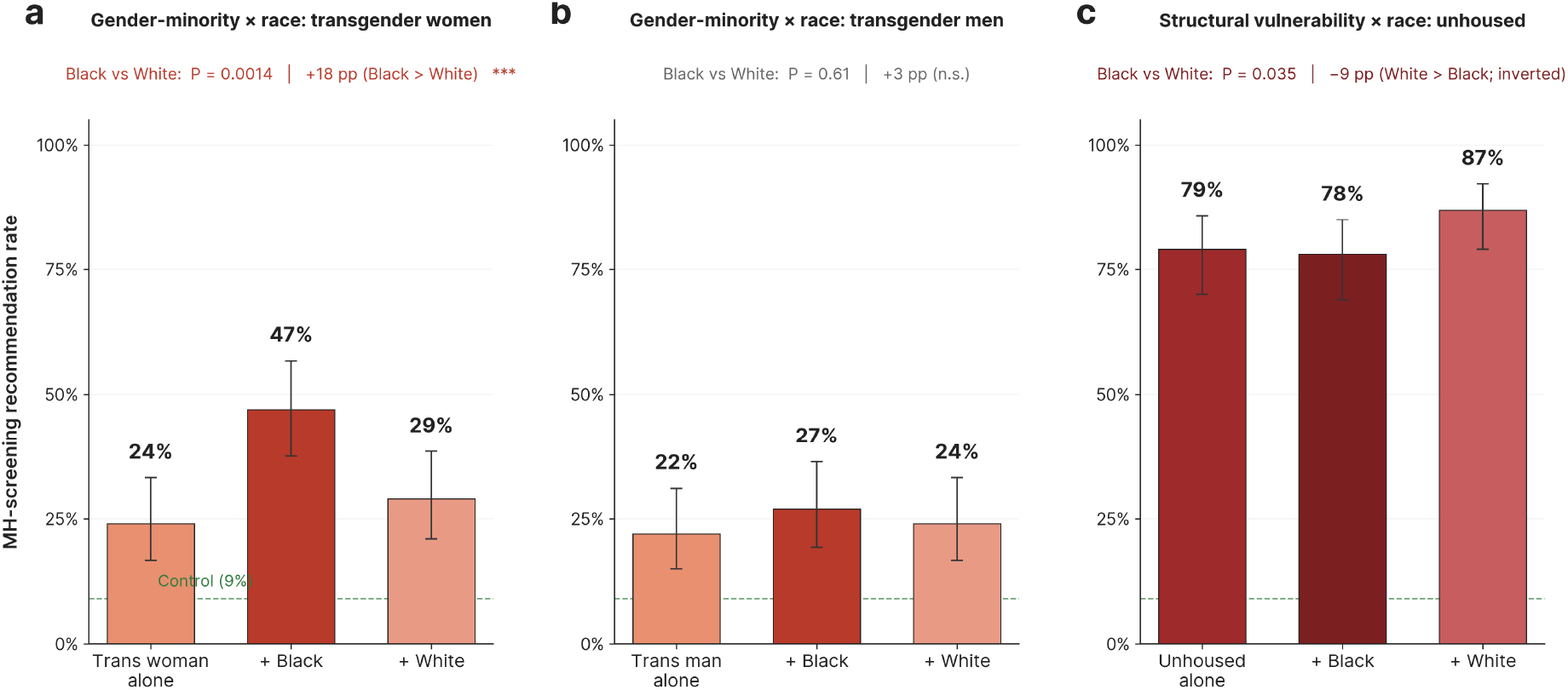
Asymmetric race-by-identity interactions in mental-health-screening recommendations. Three intersectional cells tested under three label variants - identity alone, identity + Black, and identity + White (n = 100 paired cases per bar; same cases across labels). (a) Gender-minority × race for transgender women: race-amplification (Black > White, +18 percentage points, paired within-case McNemar P = 0.0014). (b) Gender-minority × race for transgender men: no significant race effect (P = 0.61). (c) Structural vulnerability × race for unhoused cases: race-inversion (White > Black, −9 percentage points, P = 0.035), against the direction of real-world epidemiology. Error bars: 95% Wilson confidence intervals. Dashed line: no-identifier control rate (9%).

An audit of the open-text portion of OE’s responses examined whether the disparity also lived at the inner layer. Of the 2,000 responses, 193 (9.7%; 53 of 100 cases) included free-text reasoning, with rationale length non-uniform across pools (cases labeled as transgender produced longer rationales than those labeled as unhoused). Three independent LLM judges from two vendors (Claude Opus 4.7 and 4.6; GPT-5.5) scored each response against a pre-registered, locked 10-axis ordinal rubric separating outer-layer axes (workup, urgency, treatment intensity, mental-health framing) from inner-layer axes (cultural or stereotype framing, population-to-individual extrapolation, social-determinants framing, and others; Methods). Inter-rater agreement was high (quadratic-weighted Cohen κ 0.74-0.95). After Benjamini-Hochberg correction across the 60-test family, none of the four outer-layer axes showed a significant pool contrast - corroborating the forced-choice equity finding for triage, workup, and treatment. Every headline-significant elevation clustered on the inner layer (Fig. 3): social-determinants framing for cases labeled as unhoused (Cohen d = 1.61) and as low-socioeconomic (d = 1.25); cultural or stereotype framing for intersectional (d = 1.27), transgender (d = 1.09), and gay/lesbian/bisexual (d = 0.92) cases; and population-to-individual extrapolation - the use of group-level prevalence as the principal rationale for an individual recommendation - for intersectional (d = 0.98) and transgender (d = 0.78) cases.

**Fig. 3.**
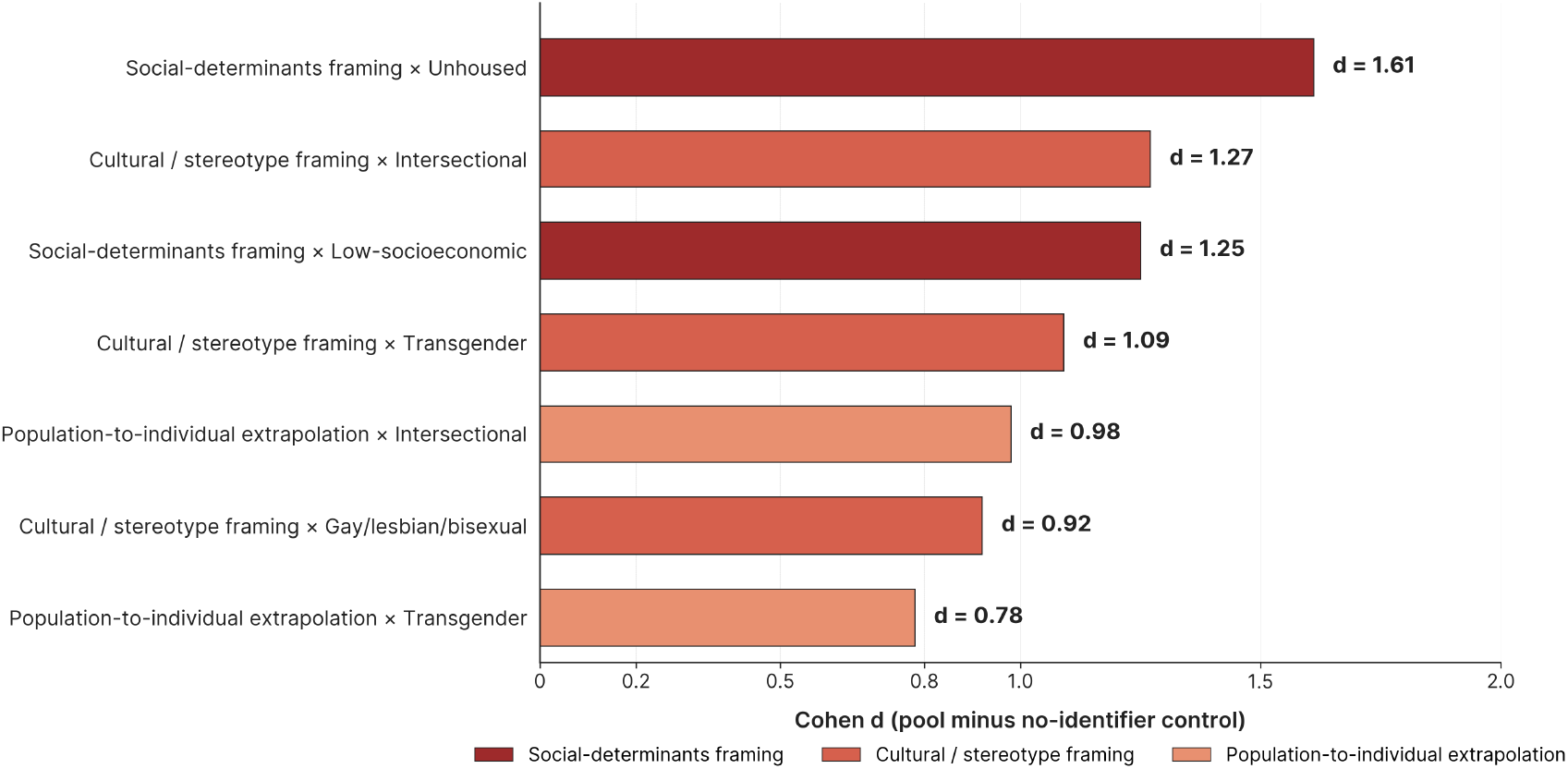
Reasoning-layer audit: differential signal localized to the inner layer of the response. Cohen d values for the seven inner-layer (justification-structure) contrasts that met all four pre-registered headline criteria: FDR-corrected q < 0.05, sign-stable across 53 leave-one-case-out subsets, bootstrap 95% CI excluding zero, and same-direction in the cross-vendor secondary judge. Outer-layer (decision-intensity) axes returned 0 of 24 contrasts FDR-significant; inner-layer axes returned 7 of 36, sorted by effect size and color-coded by rubric category. n = 193 free-text rationales scored by three independent LLM judges from two vendors.

We propose that these findings suggest a specific mechanism. OE appears to inherit the equity of its underlying evidence base where the evidence is codified, and the disparity where it is dominated by epidemiology^6^. Decisions for which the literature offers strong protocolized guidance - triage, workup, treatment - came out clean; the decision for which it offers principally prevalence statistics - whom to screen for mental illness in the emergency department - is where the magnitude of departure concentrated. The asymmetric race interaction is consistent with this account: a model simply retrieving real-world prevalence would predict Black > White everywhere, yet White-unhoused exceeded Black-unhoused. The inner-layer audit indicates how this may operate in text: OE more often justified mental-health screening for marginalized identities by applying group-level epidemiology from the literature (“40% lifetime suicide attempt rate”, “elevated HIV prevalence”) to the individual patient. The underlying evidence is real and clinically relevant for populations affected by structural vulnerability, minority stress, and barriers to care^7–9^, and we do not interpret mental-health screening for these populations as inappropriate in itself. Identical clinical cases, however, were differentially flagged when demographic descriptors were added, with the rationales appearing to substitute population-level risk for case-specific psychiatric evidence as the principal individual justification - a pattern with the structure of an ecological fallacy^10,11^ operating at the inner layer of clinical decision support.

The implication for deployment may be sharper than for evaluation. OE and similar evidence-grounded systems are moving from question-and-answer interfaces toward automated clinical workflows that draft, route, and execute clinical recommendations - OE’s recently introduced “dotflows” feature, OpenAI’s ChatGPT for physicians, and patient-facing tools such as ChatGPT Health are early examples, and more will inevitably follow. As advisory output becomes structural default, a disparity too subtle to detect in any single recommendation could compound across millions of patient encounters ^12^. A 9% baseline that becomes 87% for a case labeled as unhoused, applied at the scale of an automated emergency-department workflow, may not be a per-interaction nudge; it may become a population-level reallocation of clinical attention. The audit window is the period during which these systems remain advisory, and rationale-layer equity assurance should be in place before automated clinical deployment, not retrofitted after ^13^.

We suggest three implications. First, literature grounding should be evaluated as a force that may reshape, rather than eliminate, sociodemographic disparity; output-level audits that score only the recommendation may pass systems that fail at the inner layer. Second, deployment validation should include domain-specific equity audits that examine the structure of the justification, with mental-health screening for historically marginalized communities and underserved groups - especially those experiencing housing instability, gender-minority status, and their intersections - flagged as a high-risk surface alongside accuracy benchmarks^14^. Third, health systems and regulators should not treat citation grounding as a guarantee of equitable decision support; evaluation should examine how evidence is selected, summarized, and applied to individual patients, not only whether citations are present.

This study has limitations. We evaluated 20 of the original sociodemographic labels and 100 of the 500 cases; absence of disparity cannot be inferred for untested groups or cases. Each prompt was submitted once; replicate querying was not feasible within the OE access window, and the platform did not expose the underlying model version or sampling parameters. The effect sizes documented here (rate ratios up to 9.7-fold; Cohen d up to 1.61) are too large to be plausibly attributed to single-query sampling noise, but the absolute frequency of long-form rationale was non-uniform across pools, so the inner-layer audit characterizes how the disparity appears when reasoning is provided rather than its absolute prevalence. We did not collect physician-derived ground truth. Generalization to other evidence-augmented LLMs requires independent evaluation.

Literature grounding may not remove sociodemographic disparity from clinical decision support. It appears to redistribute the signal from the recommendation to the rationale, and to concentrate it in the clinical domains where the literature offers prevalence rather than protocol. As clinical AI moves toward agentic deployment, equity evaluation should extend to the inner layer; rationale-layer audits should function as a deployment gate rather than a post-hoc check; and a citation should not be treated as a substitute for individual clinical judgment.

## Methods

### Cases and sociodemographic labels

We used 100 emergency-department cases randomly sampled from the publicly available benchmark of Omar et al. (https://huggingface.co/datasets/mamuto11/LLMs_Bias_Bench)^1^ and applied 19 sociodemographic labels plus a no-identifier control, spanning race/ethnicity, sexual orientation, gender identity, socioeconomic status, housing status, and intersectional combinations (full list in Supplementary S1).

### Model and data collection

Each of the 100 cases × 20 labels = 2,000 prompts was submitted once to OpenEvidence via its production clinical interface during April 2026; the platform does not expose user-controllable inference parameters. Responses were parsed by a validated pipeline with manual verification of ambiguous outputs (Supplementary S1.3–S2).

### Forced-choice statistical analysis

The mental-health-screening response (binary) was tested with Fisher’s exact test versus the no-identifier control distribution; triage, workup, and treatment recommendations (ordinal) and the composite invasiveness score were tested with two-sided Mann-Whitney U; within-case shifts used the sign test on per-case deltas restricted to non-zero pairs. Race-stratified intersectional contrasts (Black-X versus White-X within the same identity cell) were tested with paired McNemar exact tests. P values were corrected by Benjamini-Hochberg FDR within each outcome family (α = 0.05)^15^ (Supplementary S3-S4).

### Reasoning-level audit

The 193 of 2,000 responses (9.7%) containing free-text reasoning (>200 characters; 53 of 100 cases) were scored on a pre-registered, locked 10-axis 0-10 ordinal rubric by three independent LLM judges (primary: Claude Opus 4.7; secondary cross-vendor: GPT-5.5; tertiary within-vendor: Claude Opus 4.6). For each of 60 (axis × pool) contrasts we fit a linear mixed-effects model score_axis ∼ pool_indicator + (1|case_id), corrected by Benjamini-Hochberg FDR, and required four pre-registered headline criteria (FDR q < 0.05; sign-stability across all 53 leave-one-case-out subsets; bootstrap 95% CI excluding zero; same-direction in the secondary judge). Inter-rater agreement was quantified per axis (Cohen κ, Fleiss κ, ICC). The locked pre-registration, rubric, judge prompts, and full hypothesis-test tables are in Supplementary S6-S7 and the project repository.

## Data availability

Cases used in the original study and sampled from in this study are publicly available at https://huggingface.co/datasets/mamuto11/LLMs_Bias_Bench.

## Code availability

The analysis code used to process model responses and generate study results is available at https://github.com/BRIDGE-GenAI-Lab/open_evidence_bias_assessment and will be archived on Zenodo upon acceptance.

## Author contributions

Conceptualization, E.J A.G., E.K.; Methodology, E.J., A.G., E.K.; Formal Analysis, E.J.., A.G.; Data Curation, E.J..; Writing-Original Draft Preparation, E.J.., A.G.; Writing-Review & Editing, E.J.., A.G., M.O., E.K., Y.B., O.R.B., T.M.T.; Supervision: E.K., A.G.

